# Retrospective Study for Safer Management for Citizen’s Marathon: A Medical Support Perspective

**DOI:** 10.1101/2025.02.05.25321727

**Authors:** Fumihiro Ogawa, Riichiro Nakayama, Yusuke Nakayama, Yoji Yuasa, Tomohiro Kamagata, Kohei Takahashi, Ryosuke Furuya, Shohei Imaki, Ichiro Takeuchi, Yokohama Marathon Management Committee

**Author notes:** Corresponding author: Fumihiro Ogawa, MD, PhD Department of Emergency Medicine, Yokohama City University, School of Medicine 3-9 Fukuura, Kanazawa-ku, Yokohama, Kanagawa, 236-0004, Japan.

## Abstract

**Background:** Although the number of individuals who enjoy sports every day is increasing owing to growing health consciousness, marathons pose health risks and can cause various types of health damage, including cardiac arrest. Furthermore, it is important to establish a system for predicting dangers and providing medical care in advance, including securing routes for transporting individuals in need of medical care during mass-gathering events.

**Objective:** This study aimed to investigate cases requiring medical care that occurred since the introduction of the full marathon at the Yokohama Citizen’s Marathon in 2015 and to examine the medical care system and its safety in relation to efforts to build a safer medical system for large-scale marathon races.

**Methods:** We retrospectively examined the incidence of health disorders and cardiac arrest during the Yokohama marathon race using the medical records provided by our committee from 2015 to 2024. Additionally, we retrospectively examined the changes in the medical support system.

**Results:** The total number of participants in the full marathon was 134,946 (total number of runners, 147,861), while the number of medical staff was 4,669 (total number of runners, 3.1±0.4%). There were 136 cases of emergency transport (0.1%), of which 27 required hospitalization (0.02%), and 3 cases of cardiac arrest during the competition (0.002%). All patients were quickly resuscitated on-site with cardiopulmonary resuscitation, and no deaths occurred. The patient presentation ratio (PPR), which represents the number of injuries and sickness per 1,000 runners, was 14.76 (range, 11.1–17.7), and the transport-to-hospital ratio (TTHR), which represents the number of emergency transports per 1,000 runners, was 0.96 (range, 0.4–2.1).

**Discussion:** Although the number of emergency transports increased and decreased owing to changes in the timing of the event and changes in temperature, it was possible to provide faster and more appropriate medical care to those in need of medical assistance. Analyzing these results annually and reviewing countermeasures to improve the medical support manual will lead to safer marathons. Such efforts will directly lead to a reduction in the number of emergency transports. In fact, the PPR was significantly lower in the current study than that in previous studies, while the TTHR was lower or similar to that in a previous study.

**Conclusion:** The Yokohama marathon is one of the largest marathons in Japan, and this study identified some participants requiring care on-course every year. Analysis of annual injury data has enabled better prediction and response to injuries, leading to safer marathon events. Most medical complaints were minor illness caused by dehydration or musculoskeletal in nature; however, there were life-threatening conditions such as CA that highlight the need for detailed planning, multi-disciplined coordination, and communication to ensure a safe and secure event. More detailed analysis of past race data under various environmental conditions is essential for running a safer large-scale citizen marathon, and the establishment of “All Yokohama” medical support system that includes local medical institutions, volunteer staff, fire departments, police, and medical staff contributes to running a safer citizen marathon.

## Introduction

Running has been popular since the 1970s (1), but the number of runners and running events has steadily increased since 2000 (1), as running is one of the most efficient ways to build physical fitness and is often associated with longevity (1). Running among adults is one of the most popular physical activities worldwide, and many cities in Western societies have their own recreational running events. Similarly, the number of marathons and road races in Japan has increased (2).Given the number of participants on this scale, marathon events fall into the “mass-participation events” category. This is traditionally defined as an event with more than 1,000 participants who gathered at a specific location at a particular time (3). Mass-gathering events have an impact on the local healthcare systems that host these events. Because of the concentration of runners and spectators at these events, medical resources can be overwhelmed if large numbers of people become injured or sick as a result of participation, which can delay public safety response to medical emergencies in the broader community (4). If an event is overly dependent on local emergency medical services (EMS), the ability to serve the public may be compromised and the host community may not receive needed medical care (5, 6). Therefore, medical planners for marathons and similar mass-participation events need to provide care that does not rely solely on local EMSs and hospital systems to determine the extent of medical care needed by event participants (7). While many injuries at mass-gathering events are minor, underscoring the importance of first aid (FA) stations and FA personnel at these events, some mass-gathering events also increase the serious traumatic injuries or illnesses (8–10). In the latter case, medical care should be provided in a two-tiered system: emergency care to respond to serious injuries and on-site care to treat minor complaints (6). Finally, it must be recognized that mass-casualty incidents may involve disasters or incidents with large numbers of casualties. Therefore, on-site healthcare systems must also be prepared to conduct emergency response activities and implement disaster response plans (7, 11). A healthcare system that can meet these demands will reduce the overall burden on local EMSs and hospital systems, improve the quality of care provided to event participants, and ensure that the public is not adversely affected by the presence of a mass-gathering event in the community.

Mass-gathering events were suspended or scaled down worldwide in 2020 due to the spread of coronavirus disease 2019 (COVID-19), but now that the COVID-19 pandemic has subsided, such events are being held as usual. As described above, marathon running is extremely beneficial for physical fitness. However, there are concerns that it may cause serious health problems, including cardiac arrest (CA) during races (12). In a previous report, the incidence rate of CA during marathons in Japan was 2.18 per 100,000 participants (12).

In a previous report, the incidence of CA during marathon races in Japan was 2.18 per 100,000 participants (12). To respond to emergencies, including CA, during a marathon race, it is important to establish a large-scale medical control system, such as preparing mobile automated external defibrillator (AED) units to provide rapid cardiopulmonary resuscitation (CPR) for on-site response and emergency transport after on-site response. Regarding the response to injured or sick runners, excluding CA runners, it is important to establish a medical rescue system in advance, considering weather, temperature, and other factors to ensure safety. In addition, it is extremely important to establish a system for predicting danger and providing relief in advance during mass-gathering events, including securing routes for medical care and transportation of individuals in need of relief.

In this study, we aimed to discuss medical safety management for marathon races and examine their problems and prospects.

### Objectives

This study aimed to investigate cases requiring medical care that occurred since the introduction of the full marathon at the Yokohama Citizen’s Marathon in 2015. Additionally, it aimed to examine the medical care system and its safety, contributing to building a safer medical system for large-scale marathon races. The purposes of this study were (1) to determine the adequacy of medical intervention for CA runners with uncertain risk factors for CA on a marathon course; (2) to reduce the burden on surrounding medical institutions in the event of a large number of injured or sick runners, including CA runners; and (3) to determine the usefulness of past data analysis for mass-gathering events, such as a full marathon.

## Methods

We retrospectively examined the incidence of health disorders and CA in citizens during the Yokohama marathon race using the medical records during the marathon races provided by our committee from 2015 to 2024. Additionally, we retrospectively examined the changes in the medical support system.

### The Characteristics of the Yokohama Marathon Course

The Yokohama Marathon Course is unique, covering 42.195 km through the bay area of Yokohama City, Japan. In addition, the race experience is affected by weather conditions. On sunny days, the direct sunrays are strong, increasing the perceived temperature; however, if the weather is bad, the perceived temperature decreases. The 15.5 km course includes steep banks, along a breezy highway with no audience, making it extremely challenging for runners.

### Medical Support Staff and System

The medical support staff consists of doctors, nurses, and paramedics from emergency and intensive care departments, or local medical clinics who are familiar with the local area. Yokohama City is responsible for gathering information and assembling the minimum necessary number of medical personnel. Medical trainers from each institute cooperate to form a medical support team.

The medical support system included 12 FA stations positioned every 3–5 km on the marathon course, an FA station after the finish line, and a temporary clinic for medical intervention outside the marathon course. FA stations and temporary clinics are staffed with medical personnel, including physicians, nurses, and medical students. These stations and clinics are equipped with basic life support (BLS) and FA equipment. Besides, the temporary clinics are equipped to perform medical interventions, including infusion and intubation, among others. All medical staff initiate FA and advanced life-saving procedures on patients who enter on their own or are brought in by other medical support teams or race officials.

### Basic Life Support and First Responder Teams with Rapid Mobile Automated External Defibrillator System

The rapid mobile AED system aims to perform immediate bystander CA and defibrillation in the event of CA during marathons. The teams were named the BLS team before the pandemic and the First Responder (FR) team after the COVID-19 pandemic. Therefore, we positioned these teams along the roadside locations and used a global positioning system to pinpoint their exact location, intending to initiate CPR within 1 min and give an AED shock within 3 min. These staff members are trained in standard-performance CPR as medical doctors, nurses, and paramedics in Japan. They also perform field triage and emergency care during heatstroke, shock, trauma, and medical emergencies. As shown in Figure 1, from the BLS team before the COVID-19 pandemic (until 2019), to theoretically ensure that an AED could reach the CA runner within 3 min, one BLS team leader was positioned every 1,000 m along the marathon course (a total of 42 leaders) and a BLS member (approximately 450–500 members) was positioned every 80 m. After COVID-19 (since 2022), this was the same theory as the BLS team, but owing to personnel allocation, the policy was to place 49 FR team leaders on the marathon course, with each FR team leader assigned four team members (a total of five persons), and, as part of the emergency action plan, a rapid mobile AED system will be built and operated by pairing teams on bicycles to treat runners who initiate resuscitation in CA cases by various types of medical personnel.

**Figure 1.**
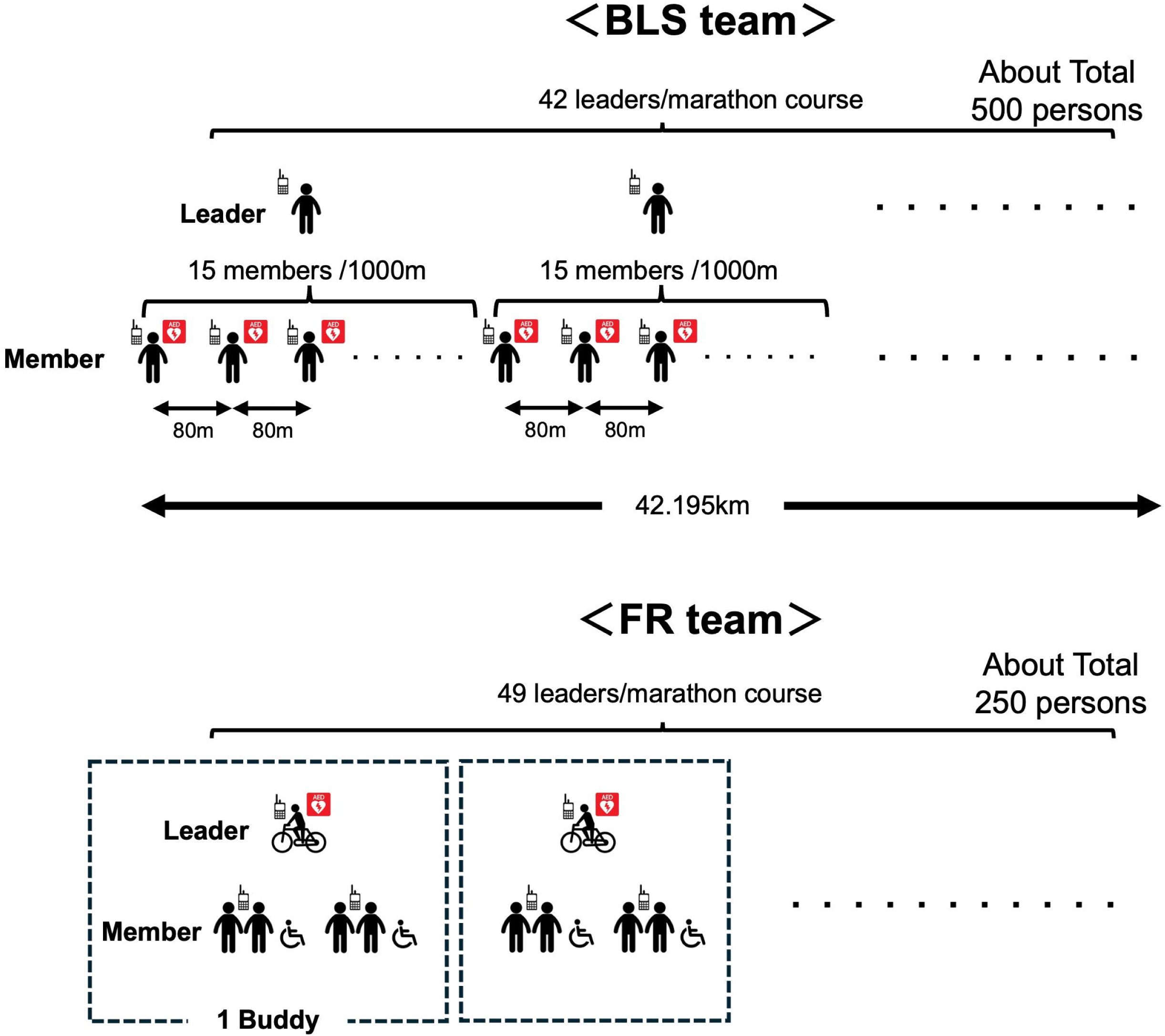
BLS and FR team System Before COVID-19, one BLS team leader was positioned every 1,000 m along the marathon course (a total of 42 leaders), and a BLS member (approximately 450-500 members) was positioned every 80m. After COVID-19, this is the same theory as the BLS team, but due to personnel allocation, the policy was to place 49 FR team leaders on the marathon course, with each FR team leader assigned four team members (total 5 persons), and to operate the team in buddy pairs with bicycle created the rapid mobile AED system to treat runners. BLS: basic life support, FR: first responder, COVID-19; corona virus infection disease 2019, AED: Automated External Defibrator

### Roadside Volunteers

Volunteers are stationed everywhere with the management manual. They are non-medical personnel trained in FA and BLS. In the event of an emergency, they call the headquarters, informing them of the location and situation, and activate the BLS or FR team.

### Running Physicians

Physicians, nurses, and paramedics run side-by-side with participants during the race. They evaluate the patient’s condition and notify the dispatch center to activate a BLS team, FR team, or command control center via a cell phone. The placement of running physicians depends on the completion times of individual marathons, and similar to regular marathon runners, they start in seven blocks based on their completion times. In addition, running physicians who participate for the first time are placed in the final block to maintain balance. Each running physician is instructed in advance to keep a minimum of 100 m between each other and to participate in the marathon.

### Dispatch and Medical Command Control

At the command control center, a physician acts as the medical director, who is responsible for medical support, and some physicians act as the medical controller between the command center, FA center, and temporary clinic via phone or transceivers at any time. Moreover, approximately 20 paramedics act as dispatchers in the fire office departments. In cases of CA, injury, or sickness, the command control center gets some information from bystanders or physicians at the FA place who witness and perform basic telecommunication. The medical control doctor provides advice via phone in the case of an emergency. If an incident occurs on any marathon course at the command center, the medical control doctor and fire department work together to dispatch ambulances to the scene in an appropriate manner and decide the destination in advance to ensure smooth transport of the CA, injured, or sick runner. Through bystander interviews, medical records, and event audits, they create timeline data and medical records.

### Cardiac Arrest Response

If someone meets a CA case, anyone who witnesses a collapse can call a dispatcher. The dispatch unit activates the BLS or FR team. To identify a patient’s location, medical personnel, and mobile AED team at a nearby FA station, using GPS or transceivers. The team is positioned to quickly initiate FA, BLS, and defibrillation. However, multiple individuals with different roles, such as paramedics, paramedic students, running physicians, medical personnel from FA stations, BLS teams, FR teams, volunteers, and runners, can be FRs. Therefore, in this study, the person who first reached the patient and initiated chest compressions for the CA runner was considered the FR. The BLS and FR teams cannot transport patients; therefore, they call an ambulance if a patient requires further assistance. The team remains at the location until the patient recovers or the fire-based ambulance crew arrives.

## Results

### Weather and Temperature

Figure 2 shows the weather and temperature changes on the days of the marathons. The weather in 2015 and 2016 was cloudy with sunny spells, with maximum temperatures of 13.2 °C and 10.9 °C, respectively, and minimum temperatures of 6.2 °C and 4.6 °C, respectively (average temperatures were 9.4 °C and 7.3 °C, respectively). In 2017, the event was canceled owing to poor weather and conditions, and in 2018, it was rescheduled from the end of March to the end of October. Since 2018, the maximum temperature has exceeded 20 °C, and the minimum temperature has exceeded 10 °C, which may have made it more difficult for runners than before. There were no significant changes in the maximum and minimum humidity.

**Figure 2.**
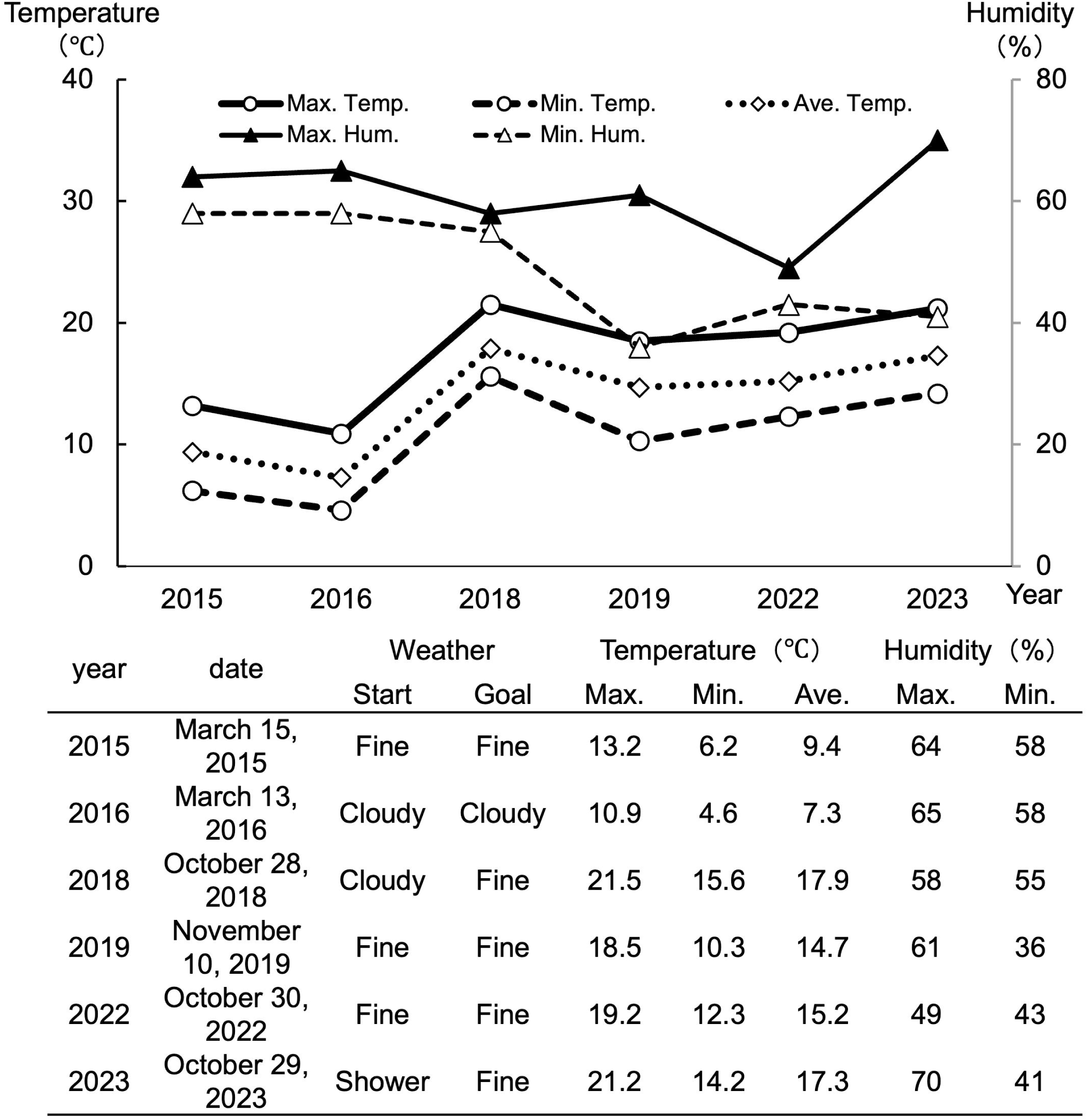
Weather and Temperature To show the weather and temperature on the day of the marathon. Temperature (maximum, minimum, average) and humidity (maximum, minimum) are displayed over time.

### The Characteristics of Runners and Medical Support Staff

The characteristics of the full marathon runners are shown in Figures 3 and 4.

**Figure 3.**
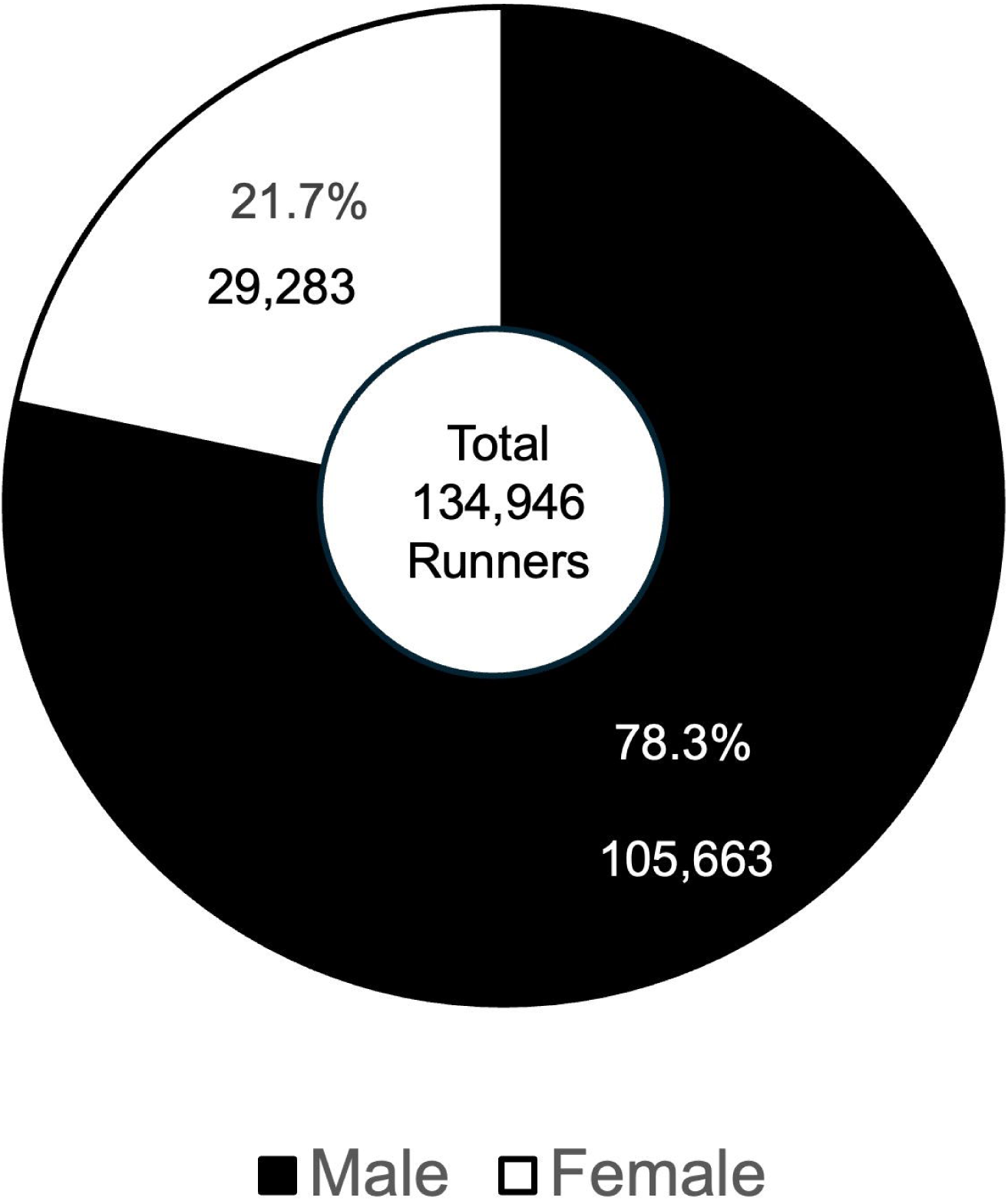
Gender Ratio of Full Marathon Participating Runners Male made up 78% of all participating full marathon runners.

**Figure 4.**
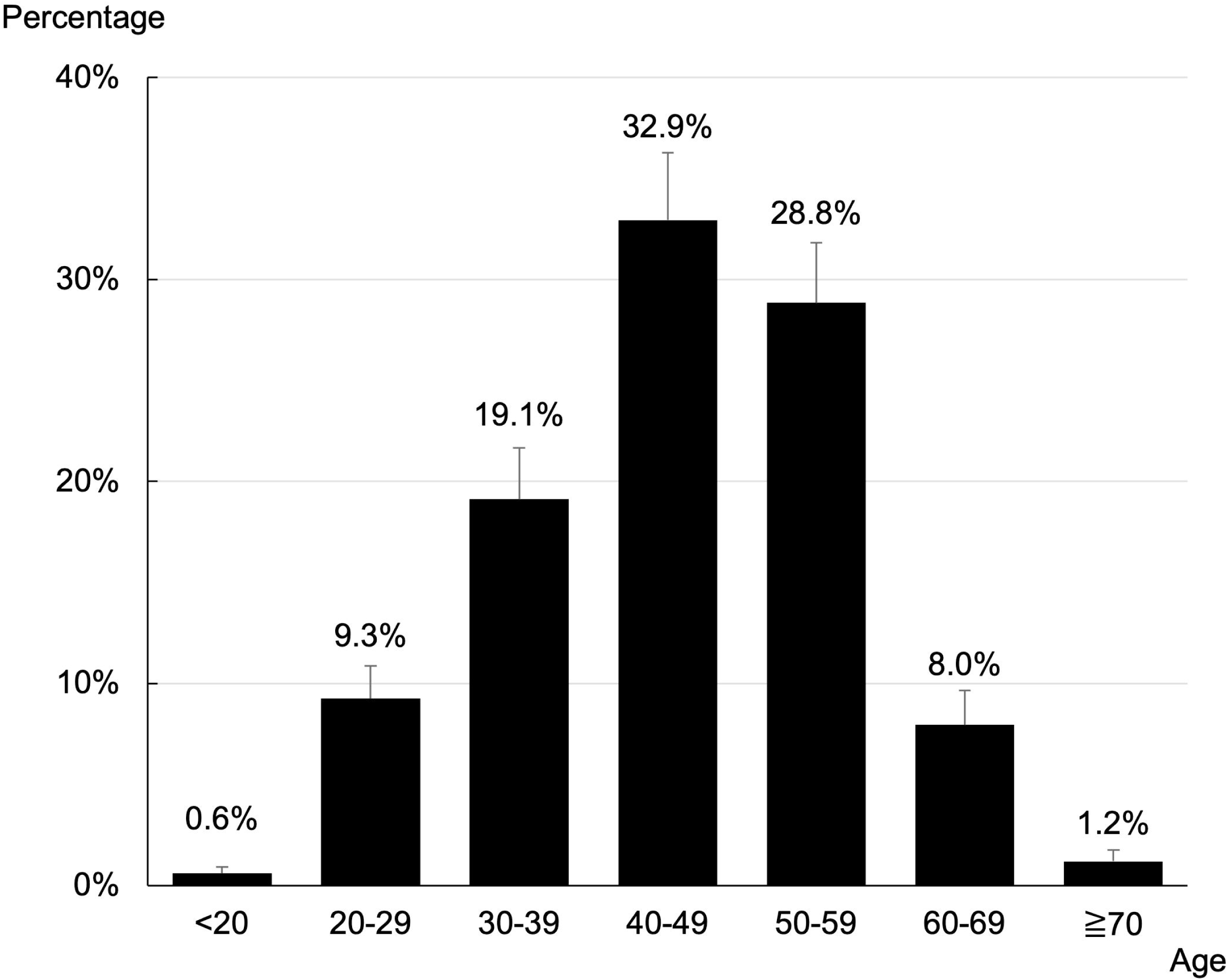
Breakdown of Full Marathon Participating Runners by Age Of all participating full marathon runners, those in their 40s and 50s account for over 60%.

Based on sex, 78% of the approximately 135,000 runners were male. Based on age, 0.6% were aged < 20 years, 9.3% were in their 20s, 19.1% were in their 30s, 32.9% were in their 40s, 28.8% were in their 50s, 8.0% were in their 60s, and 1.2% were aged > 70 years.

Therefore, the majority of participants were in their 40s or 50s.

Figure 5 shows the number of full marathon runners and their completion ratios. In the pre-COVID-19 pandemic phase (2015–2019), the number of participants in full marathons increased annually, with approximately 25,000 participants each year. The completion rate was high at 92–96%. Because of the COVID-19 pandemic, the event itself was not held between 2020 and 2022 and resumed in 2023 but was held on a slightly smaller scale in consideration of COVID-19 prevention. The completion rate was 92–93%, which was not significantly different from that in the pre-COVID-19 phase.

**Figure 5.**
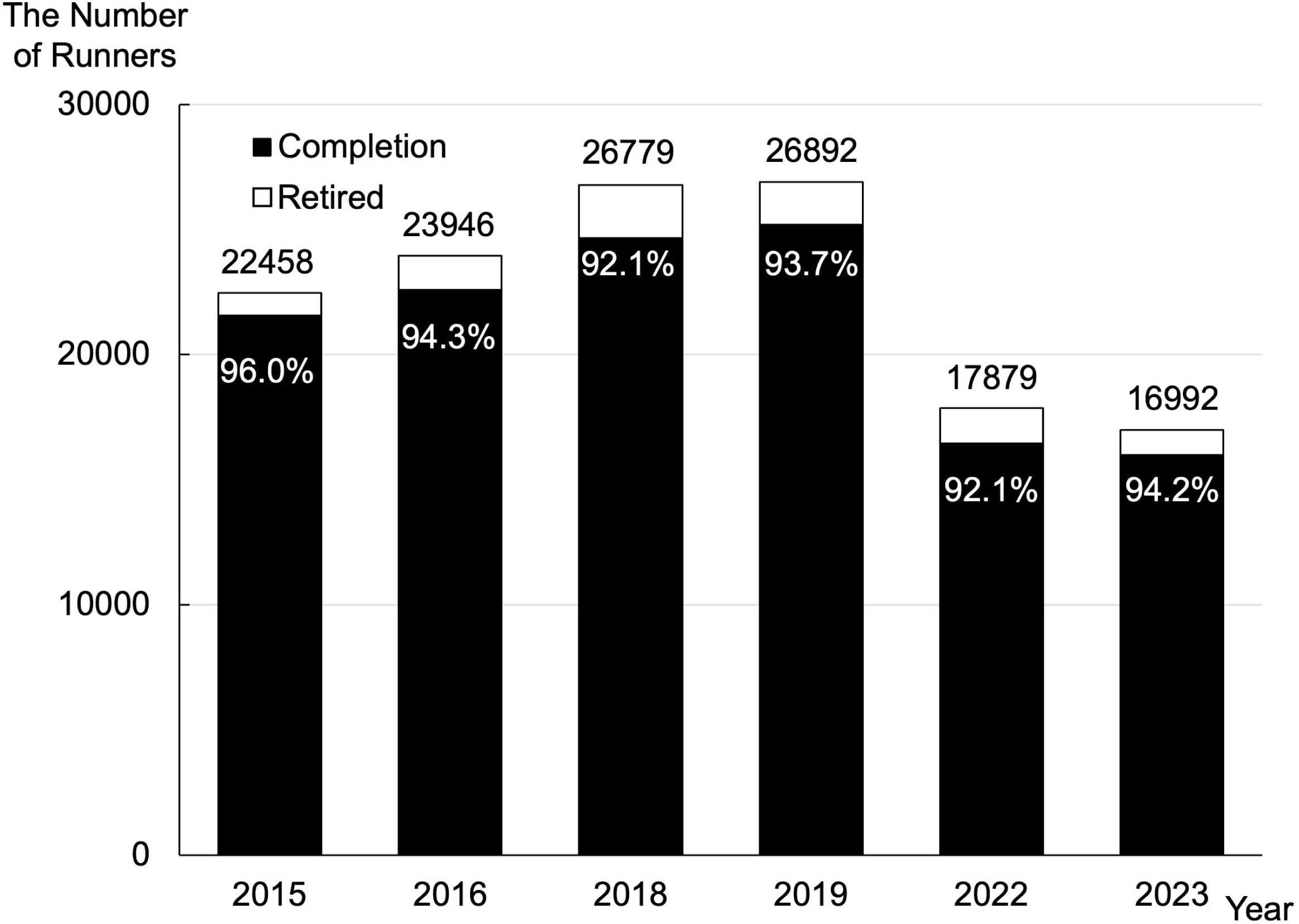
The Number of Runner and Completion Ratio To show the annual trends in full marathon participants and completion rates. In 2022, the number of applicants decreased due to COVID-19, but the completion rate did not change significantly.

In Figure 6, regarding medical support staff, the number of cooperating doctors increased annually (32–50 doctors), the number of nurses remained stable (approximately 35 nurses), and the participants of doctor runners were mixed. The number of BLS teams in the pre-COVID-19 phase increased with the change in the date of the event in 2018 (maximum 600 persons). However, in the post-COVID-19 phase, they became FR teams as a strategy to prevent the spread of COVID-19, and their numbers decreased (350–400 persons).

**Figure 6.**
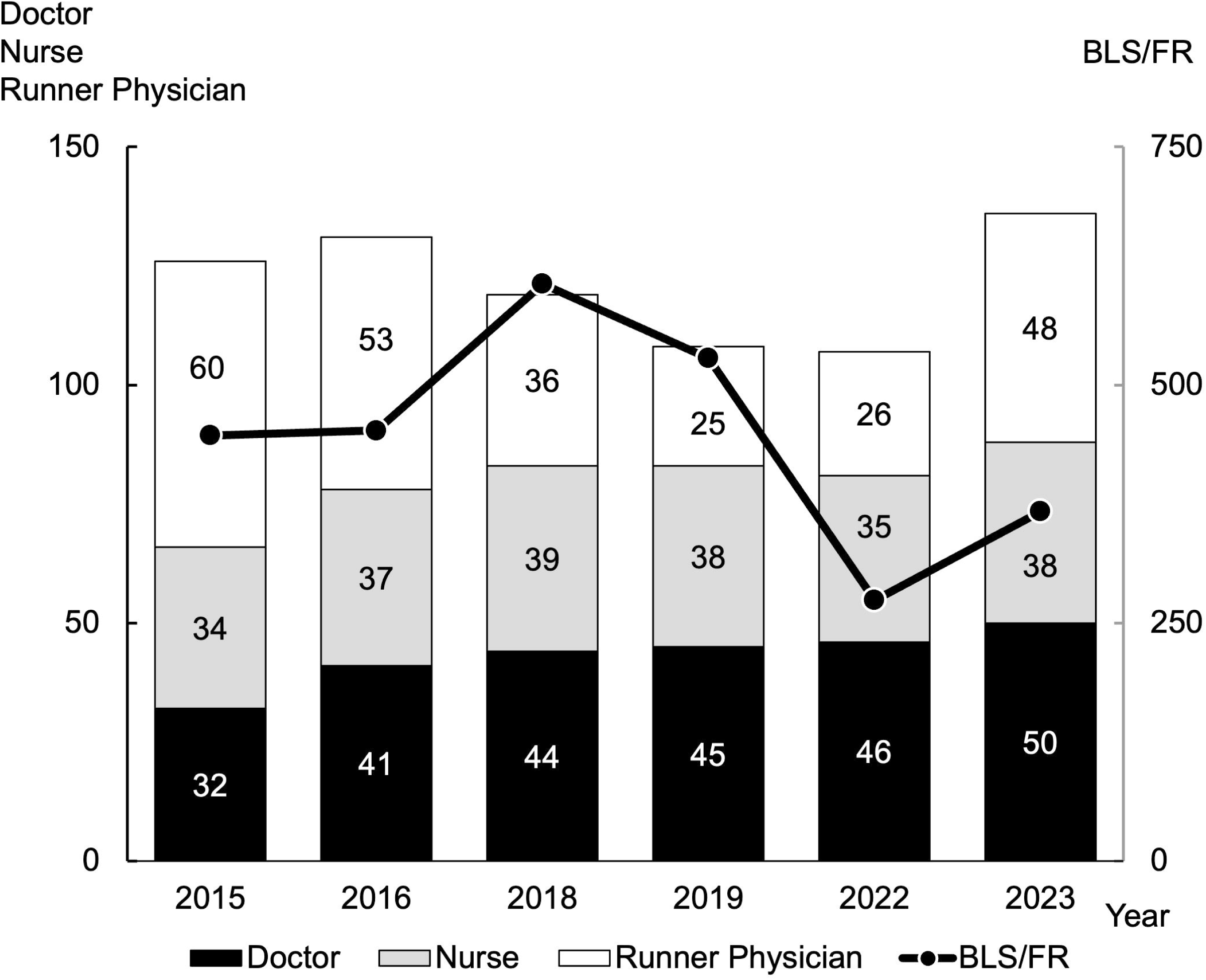
Transition of Medical Support Staff The number of medical staff at first aid stations and temporary clinics supporting the marathon has not changed much. The number of BLS/FR members have decreased for COVID-19 pandemic.

### The Number and Characteristics of Injured or Sick Runners at the First Aid Stations on the Marathon Course, after the Finish Line, and at Temporary Clinics

Figure 7 shows the breakdown of injured or sick runners at all the FA stations on the marathon course, the FA station at the goal line, and the temporary clinic outside the marathon course. In 2018, when the race date changed, the number of injured and sick runners increased across the boards (431 runners). This is thought to be due to seasonal and weather changes. As a result, in 2019, a scheme was launched to transport runners who needed medical intervention from the FA station on the course to temporary clinics as soon as possible, which led to an increase in the number of injured or sick runners treated at temporary clinics (156 runners, a total of 403 runners). There will be restrictions on Sydney participants from 2022 onward, and a decrease in injured or sick runners (316 runners in 2022 and 186 runners in 2023) was observed.

**Figure 7.**
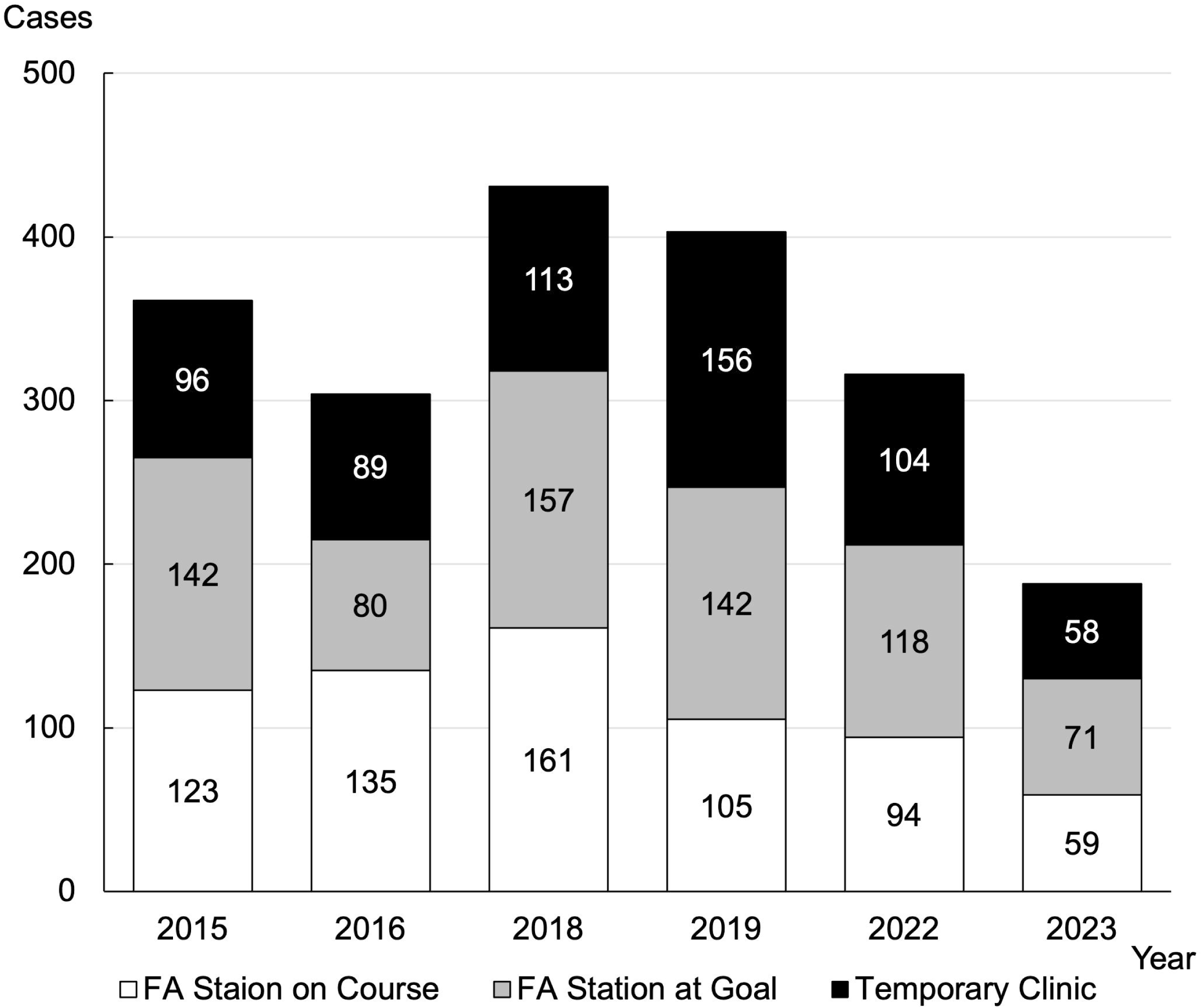
The Number of Injured or Sick Runner at the FA station and Temporary Clinic The number of injured or sick runner has increased due to the change in the date in 2018, but due to improvements in the medical support system each year, the number of injured or sick runner being treated in each department is decreasing. FA: first aid

Table 1 presents a breakdown of the symptoms of injured or sick runners, including the number of emergency transports. With a change in the timing of the event in 2018, the number of runners experiencing dehydration and heatstroke due to an increase in temperature increased (25 runners); there was also an increase in emergency transport (55 runners).

**Table 1.** Breakdown of Transport to Hospital with Their Symptoms.

| Year | Transport to Hospital | Dehydration Heat Stroke | Nausea Vomit | Seizure | Injured | Cardiac Arrest | Muscle Cramps | unknown |
| --- | --- | --- | --- | --- | --- | --- | --- | --- |
| 2015 | 9 | 4 | 1 | 0 | 0 | 0 | 4 | 0 |
| 2016 | 14 | 4 | 1 | 0 | 3 | 0 | 6 | 0 |
| 2018 | 55 | 25 | 10 | 7 | 3 | 1 | 6 | 3 |
| 2019 | 29 | 7 | 4 | 5 | 0 | 1 | 12 | 0 |
| 2022 | 20 | 8 | 1 | 1 | 5 | 1 | 4 | 0 |
| 2023 | 9 | 3 | 2 | 1 | 2 | 0 | 1 | 2 |

Figure 8 shows the number of runners transferred based on age at the FA station and temporary clinic from 2019 to 2023 (between 2015 and 2018 could not be included in these data because of uncertain details). Similar to the proportion of participating runners, most of those who were transferred to hospitals were in their 40s and 50s.

**Figure 8.**
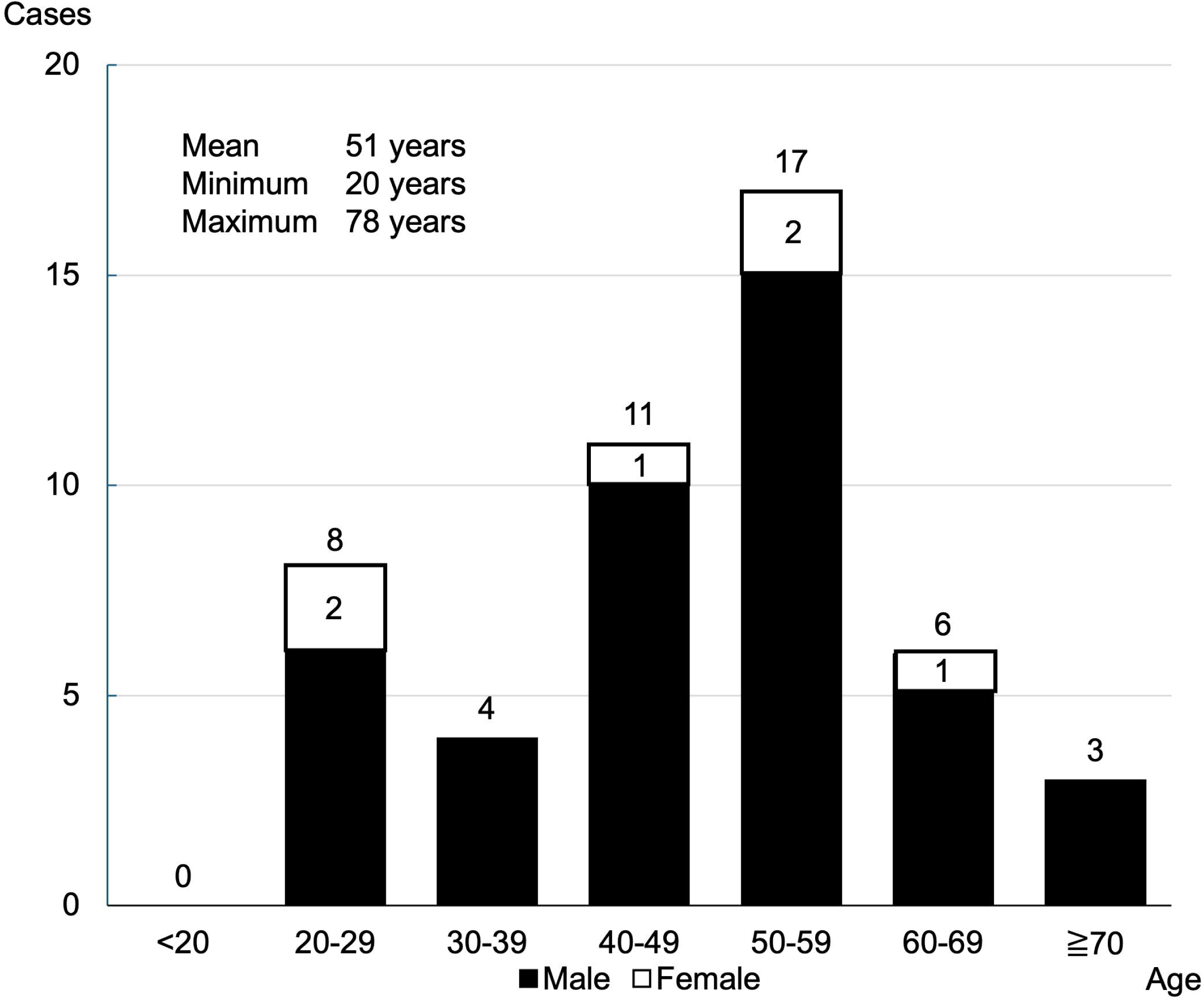
The Number of Transferred Runners by Age at the FA station and Temporary Clinic from 2019-2023 Similar to the proportion of participating runners, most of transferred to hospital were in 40s and 50s (not included due to uncertain details from 2015 to 2018). Total number is above bar graph, and the number of females is at the white bar.

Figure 9 shows the number of runners admitted to the hospital after medical interventions. Runners requiring medical intervention after emergency treatment were admitted to the hospital. Patients who required continued hospitalization were admitted because of severe dehydration (including heatstroke) and difficulty moving due to muscle cramps. In 2018, many runners were injured or sick due to heatstroke caused by the change in the timing of the event; therefore, hospitalizations were higher than those in other years.

**Figure 9.**
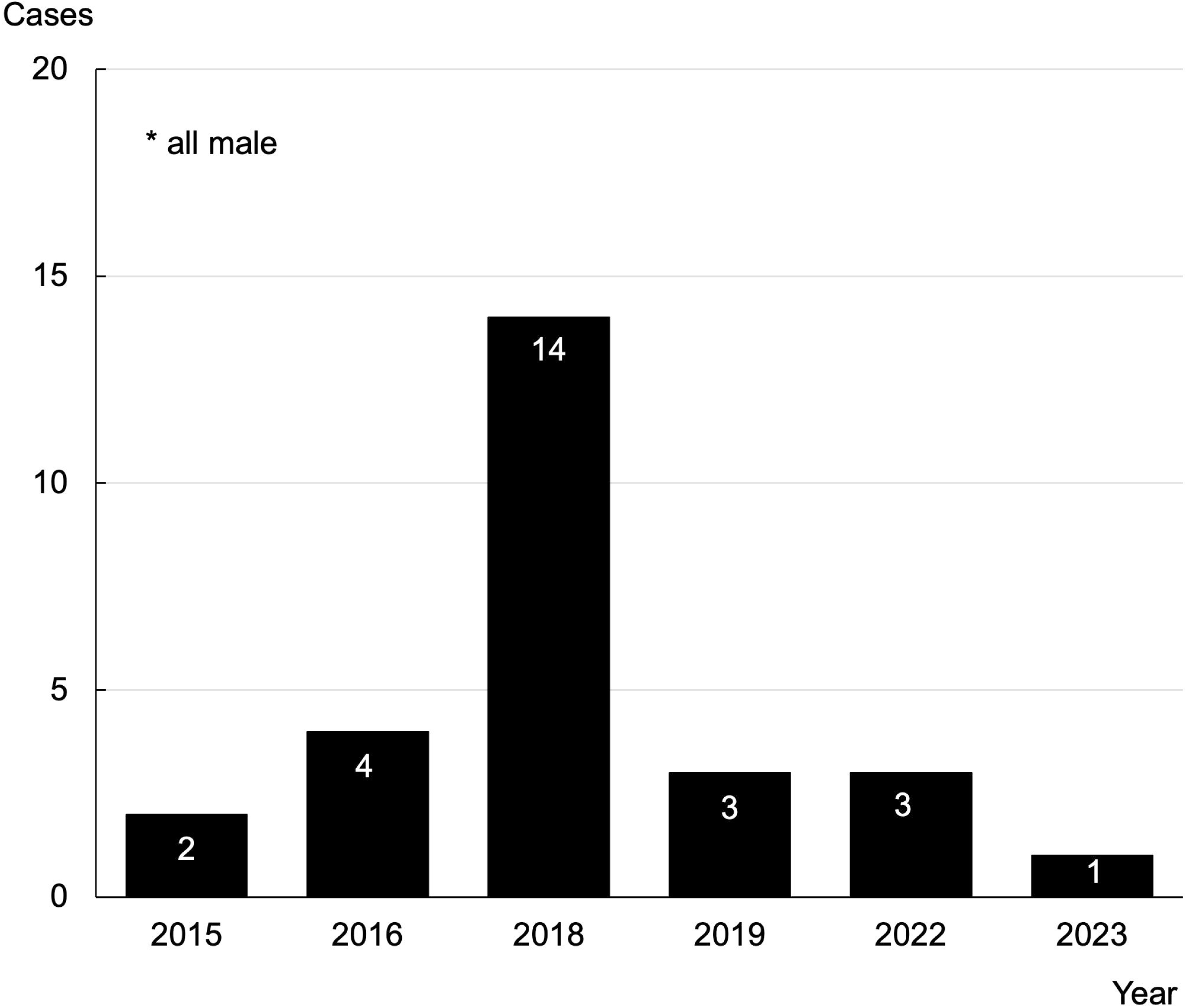
The Number of Runners Admitted to Hospital The cause of hospitalization were severe dehydration (including heat stroke) and difficulty moving due to muscle cramps. All the runners who required hospitalization were males.

### The Change in the Index of Injured and Sick Runners and Emergency Transport

Figure 10 shows the change in the index of injured and sick runners and emergency transport with patient presentation ratio (PPR) and transport-to-hospital ratio (TTHR). PPR represents the number of injuries and illnesses per 1,000 runners, and TTHR represents the number of emergency transports per 1,000 runners. The PPR was not significantly affected by completion rate, weather, or temperature (the index from 2015 to 2019 ranged from 12.7 to 16.1). However, since 2022, the coefficient may have increased because volunteers and doctor runners encouraged runners who were likely to be injured or sick to be accommodated at FA stations (17.7). In 2023, it may have been slightly affected by temperature (11.1).

**Figure 10.**
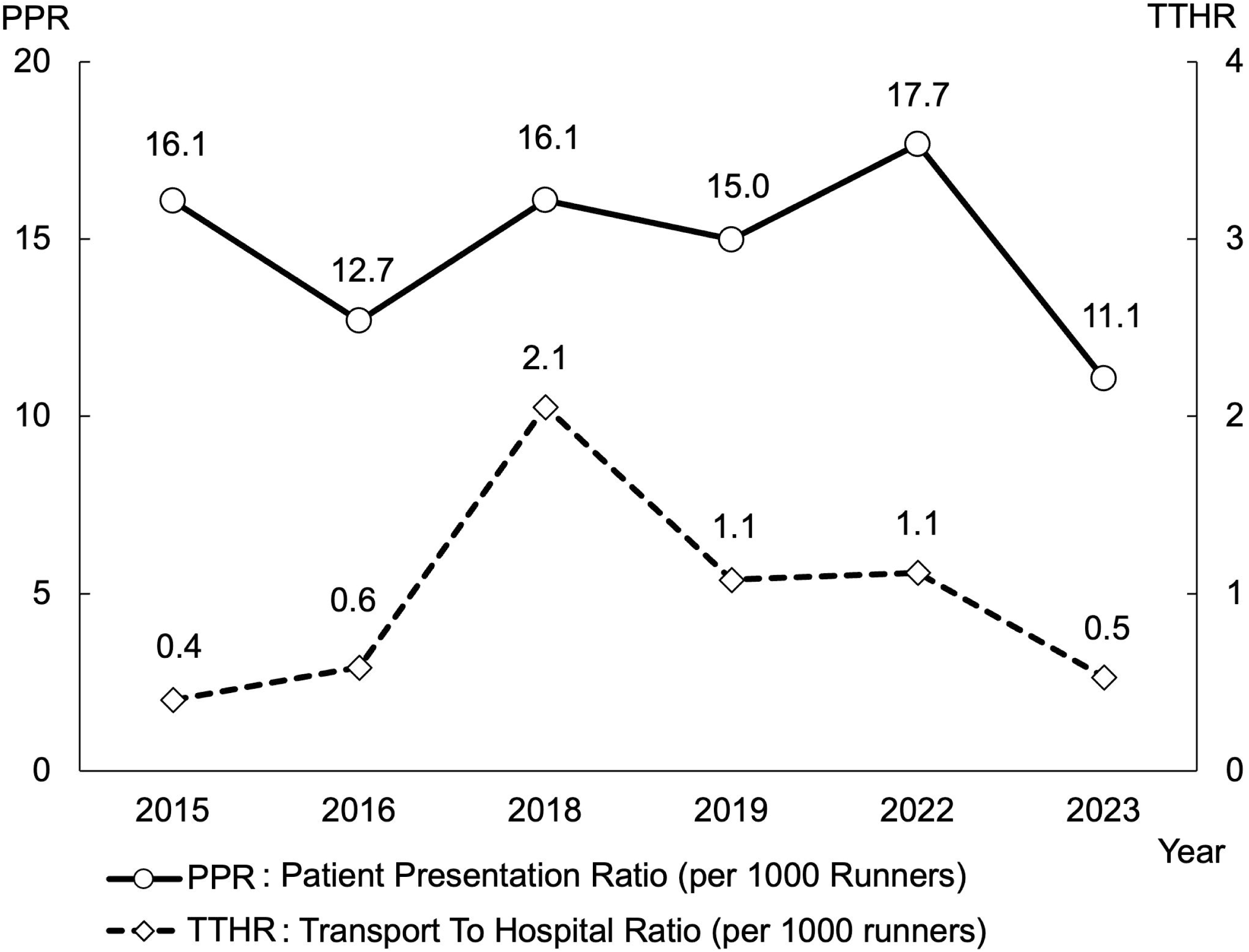
Change of the Index of Injured or Sick Runner and Runner Transferred to Hospital The trend in Patient Presentation Ratio (PPR), which indicates the prevalence of injured or sick, does not seem to have changed dramatically, but Transport To Hospital Ratio (TTHR), which indicates the rate of hospital transport, increased dramatically after the change in the date in 2018 and has since started to decrease. This downward trend may be due to improvements in the medical system through annual review.

Regarding the TTHR, the change in the timing of the marathons from March to October 2018 may have led to an increase in the number of cases of heatstroke and dehydration caused by increasing temperatures, which resulted in an increase in the number of cases of emergency transport (2.1). However, since 2019, based on the event in 2018, we have increased the number of water stations, increased the number of beds at FA stations, and strengthened triage at FA stations on the course, which has led to a decrease in TTHR because of taking measures to prevent more life-threatening injuries and sickness (1.1 to 0.5).

### Detailed Analyses of Three Cardiac Arrest Runners

Table 2 presents the details of the three CA cases that occurred during marathons.

**Table 2.**
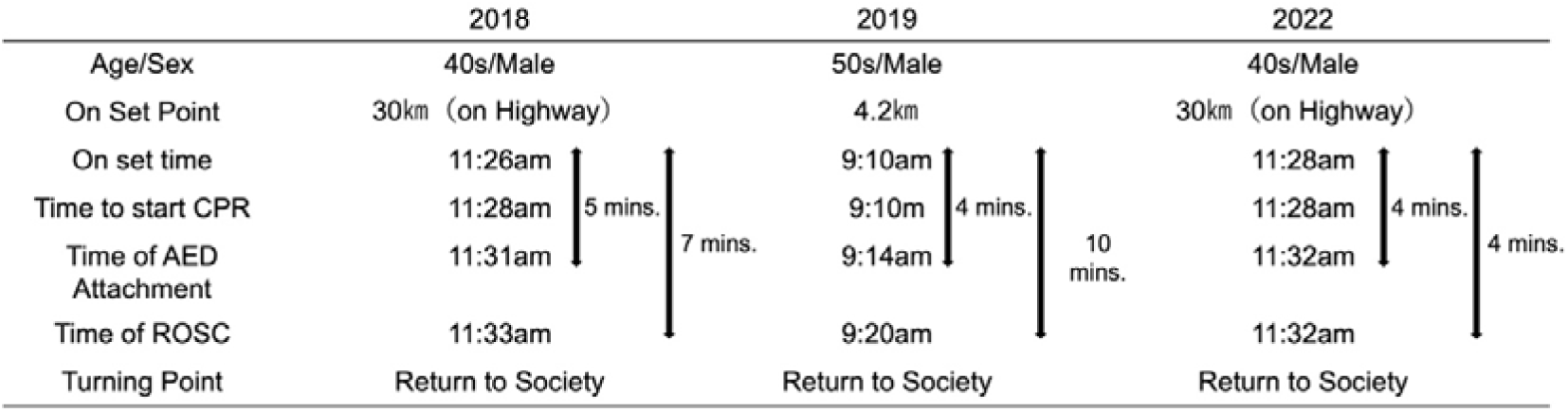
Details of Three Runners with Cardiac Arrest.

Three of 135,000 full runners experienced CA, with an incidence rate of 2.2 per 100,000. Two cases occurred approximately at the 30 km mark and one at the 4.2 km mark. All the CA runners were males in their 40s and 50s. CPR was initiated immediately after witnessing the incident, and the AED arrived within 5 min. All three patients underwent CPR at the scene and were transported to the hospital for treatment; however, they were able to return to society without any health problems. One problem is that the time taken to transport the patient from the scene to the hospital may have been delayed because of the expressway, which has limited access routes.

## Discussion

We conducted a retrospective study of large-scale citizen marathons to investigate the incidence of injuries and sickness during marathon races, runners being transported to hospitals, and cases of CA. The number of injured and sick runners and emergency transports increased, which is believed to be due to changes in weather and temperature caused by the marathons. Many of the symptoms were due to dehydration and heatstroke, and improvements to eliminate the causes through the analysis of past data may have helped reduce the number of injured and sick runners transported to emergency hospitals. The number of emergency transports was decreased by creating a scheme to strengthen triage at FA stations on the marathon course and by transporting runners to temporary clinics where medical intervention is possible. In addition, by appropriately deploying BLS or FR teams during the course, it was possible to better support runners in preventing minor injuries from being transported to emergency hospitals. In three cases of CA that occurred during the marathon course, CPA was started immediately after the incident, and an AED was attached within 5 min for all CA runners, resulting in resuscitation on the scene, and all patients were able to return to society.

### Relationship between the Environment and the Injured or Sick Runner

Regarding climate and temperature fluctuations and the increase in injured or sick runners, the change in the date of the race from the end of March to the end of October caused an increase in temperature in 2018, and there was a clear increase in the number of injured and sick runners and those who were taken to emergency hospitals. We searched for reports on the increase in temperature and the number of injuries or sicknesses during marathons using search engines, such as PubMed and MEDLINE; however, we were unable to find any research that clearly established the cutoff value for the increase in injured, sick, or CA runners. Regarding ambulance dispatches, a previous study demonstrated that the number of ambulance dispatches was the lowest when the temperature was 22.5 °C, increasing as it deviated from that point in a U-shaped relationship (13). Therefore, the relationship between exercise and the outside temperature is likely to be involved in the occurrence of injuries or sickness. Similar to dehydration and heatstroke, we improved annually to prevent these conditions by increasing the number of water stations and aid points in the marathons. Faster runners are more susceptible to the effects of weather than slower runners; among temperature, relative humidity, specific humidity, wind speed, solar radiation, and longwave radiation, the effect of temperature is the strongest and females are less susceptible to these effects than males. In our report, it is thought that injured, sick, or CA runners are more susceptible to the effects of weather than male runners (14). A previous study reported that although runners’ performance declined during rainy weather, their completion times were faster (14). In addition, the Wet Bulb Globe Temperature (WBGT), which has often been used as an indicator of heatstroke and summer exercise owing to the influence of global warming in recent years, has been reported in a study conducted on the marathon race of the Tokyo Olympics held in 2021 during the COVID-19 pandemic (15). In this study, a detailed analysis was performed using the WBGT to examine what type of heat effects will occur in outdoor sports and what countermeasures will be effective if global warming progresses. In conclusion, it was reported that it is desirable to hold safer competitions by examining the timing, time, and host country of the Summer Olympics based on WBGT trends. In our study, we also measured the WBGT annually, and as the WBGT did not correspond to the standard for restricting outdoor exercise, we did not mention it here. However, the WBGT measurements on the highway, which is the unique course design of this competition, and at the start and finish points, where indicators, such as wind speed and sunshine rate significantly fluctuate, may also have affected the number of injured or sick runners.

### Specialized Medical Support System

Regarding the response of injured, sick, or runners with CA, CA is a leading cause of mortality; a previous study indicated that 1 in 7.4 individuals died of CA (16). In adults, 39% of CAs are related to sports (16). In marathons, the cardiac death rate ranges from 0.24 to 0.39 per 100,000 runners (17, 18). A previous study reported that the incidence rate of CA was 0.54 per 100,000 runners (17). Tokyo marathon had a slightly higher rate of 0.65 per 100,000 runners than other marathon races (19). To increase the survival rate of patients with CA during marathon races, the early initiation of CPR and defibrillation with AED is important (19). In the Yokohama Marathon, 3 of 13,500 runners were confirmed to have experienced CA during the marathon race at a rate of 2.2 per 100,000. This is an extremely higher incidence rate than the previous report, but all three runners with CA were resuscitated at the scene and successfully returned to society. The most significant factor in the rescue of victims was the rapid response from the time of the incident to the use of CPR and AED. Roadside medical support that can provide initial CPR and effective defibrillation is required for big-scale sports events. Organizers of marathon races must understand the risks of CA during marathon races, and build marathon-specific medical support for delivering bystander CPR and defibrillation immediately and achieving favorable survival. Additionally, it is also important for all medical staff and volunteer staff to understand their own roles using activity protocols by themselves, and to communicate with each other. Accordingly, we propose five factors of medical support for the Yokohama Marathon. (1) proper composition and deployment of rescue and emergency response personnel, (2) efficient deployment of medical and rescue equipment, including enough AEDs, (3) preparation of protocols for communication and command/control system protocols, (4) training of emergency medical staff before the event, and (5) preparation of rescue personnel protocols and EMS operations manuals. This was largely because an emergency response manual was prepared in advance and an established response flow was shared and thoroughly understood by not only the medical rescue team but also all event staff, including event volunteers, during the briefing before the marathon. Certainly, this is not limited to CA cases; all staff members are committed to a safe medical system that quickly and accurately recognizes and evaluates the conditions of injured or sick runners. In addition, as mentioned in the medical support system, we placed the BLS and FR teams in locations to initiate CPR within 1 min and give an AED shock within 3 min with the global positioning system to determine the exact location, as early initial defibrillation is the most effective for saving life in case of CA in sports. Additionally, it is important to recognize the signs suggesting CA like gasping during marathons, as gasping is a major early indicator, observed in 90.5% of CAs (20). The BLS team before the COVID-19 pandemic until 2019, carrying FR kits and AEDs, covers every 80–100 m of the course, totaling approximately 50 staff. The FR team covered 1000 m of course, one reader, and two teams each of two staff, with a total of five staff members in a 1000-m marathon course. Therefore, there is a need to devise ways to respond to the injured, sick, and CA runners. The placement of the BLS and FR teams was based on theoretical safety calculations. In the case of a BLS team, members are placed at a certain distance (one member per 80–100 m) from each other; therefore, the activity is limited to a certain area, which has the advantage of allowing for a quick response to injured, sick, or CA runners. However, the disadvantages are that one team leader must manage 15 members, making it difficult to keep track of all members, and many medical supporters are required for placement. Here, FR team creation was considered the best way to deal with the difficulty of securing personnel during the COVID-19 pandemic. Because one team leader is paired with four members, for a total of five buddies, information sharing has improved dramatically and better communication has been achieved with transceivers, making it easier for the team to grasp the situation. Furthermore, because teams are placed in blocks, cooperation between FA stations has become easier, making on-site rescue activities more efficient. The medical team at the FA station, temporary clinic, and marathon race headquarters have formed a cooperative system regularly and are organized with close relationships between emergency doctors from surrounding institutions, clinic doctors in the area, and the local fire department so that they can easily respond to emergencies. Since the Marathon Medical Management Committee was first established, the most important thing has been to emphasize “on-site judgment,” and a medical coordinator and sub-coordinator are assigned to each FA station, so that information from the FA station can be shared with the headquarters immediately, and a system for rapid emergency transport is in place, if necessary. As mentioned above in the course characteristics of the Yokohama Marathon, the marathon course includes a highway, and because access routes in the event of an emergency are limited, there is a high possibility that normal emergency response will delay transporting injured, sick, or CA runners to the hospital. To solve this problem, ambulances, private ambulances, and hospital ambulances are stationed along the marathon course; dispatch orders are issued directly from the EMS department of the race headquarters, and the medical support team of the race headquarters searches for and decides on the destination in advance, enabling faster transport to the hospital.

### Medical Operation Manual and Annual Review System

Regarding the medical operation manual, we analyzed the problems that occurred in the previous year using a medical rescue matrix so that solutions and improvements could be immediately utilized in the following year and reflect the results. During this period, the number of injured and sick runners increased owing to the change in the date of the event in 2018 (TTHR, 2.1). We analyzed the injuries and sicknesses and found that many were caused by dehydration and muscle cramps; thus, we increased the number of water supplement stations. As there were many cases of transfer to hospitals, we strengthened triage at the scene and added a protocol to immediately transport injured and sick runners who required medical intervention to a temporary clinic, thereby reducing the number of transfers to hospitals (TTHR, 1.1). When the marathon resumed in 2022, an increase in the number of injured individuals (especially those with external injuries) was observed at FA stations (PPR, 17.7), which may have been due to a decrease in exercise habits caused by COVID-19. Running injuries mainly consist of muscle injuries, sprains, and skin lesions (11). And, running easily leads to overuse injuries of the lower back and legs (21). Although few runners were taken to hospitals, the FA stations responded by increasing the space to accept individuals with minor injuries and by setting up massage spaces in 2023 (PPR, 11.1). Focused on PPR and TTHR, the PPR (14.76 [range, 11.1–17.7]) in our study is significantly lower than those of other studies (22, 23). However, the TTHR (0.96 [range, 0.4–2.1]) in our study is low or similar to those of other studies (22, 23). This is presumably due to the successful development of the medical management system, as described above, but as this can vary depending on various factors, such as the location and timing of the event, it is not possible to state that it is, in general, an excellent medical system. However, we believe that it is commendable that the values improve in the same place and simultaneously.

Regarding the prediction of injuries or sickness that may occur during a marathon, the Yokohama Marathon Committee conducts preliminary investigations. However, as these investigations are not detailed, it is important to analyze it in-depth and identify potential causes. Identifying such factors may contribute to the development of injury prevention strategies through proper training and optimization of the training environment (24).

Furthermore, the exact causes of running injuries are diverse (25) and interact with each other (24). Risk factors for running injuries can be clustered into three components: (1) personal factors, (2) running/training factors, and (3) health- and lifestyle-related factors (25). By accurately investigating and ranking these risk factors in advance, it may be possible to predict the number of injured, sick, or CA runners. Moreover, by including factors such as weather, temperature, and WBGT, the number of injuries, sick, and CA runners could be predicted.

## Limitations

This study has some limitations. First, this study was retrospective, making it difficult to trace the details of injured, sick, and CA runners. Second, the backgrounds of injured, sick, or CA runners were based on self-reporting before the marathon race, which may not accurately reflect their actual conditions. Third, regarding the treatment of injured or sick runners, although the events were held in the same area, the medical staff was different annually; therefore, there may be slight differences in the provided treatment. Finally, the results reported in this study cannot be generalized to other marathons, as they are influenced by factors such as course design, environmental conditions, duration of the event, and participant demographics. Additional multi-site studies are needed to compare marathon events, and more in-depth research is needed to assess the relationship between weather conditions, course design, volunteer and medical staff, and patient care and outcomes.

## Conclusions

With an increase in health consciousness through daily exercise habits, routine running and participation in large-scale marathons have increased. The Yokohama marathon is one of the largest marathons in Japan, and this study identified some participants requiring care on-course every year. Analysis of annual injury data has enabled better prediction and response to injuries, leading to safer marathon events. Most medical complaints were minor illnesses caused by dehydration or musculoskeletal in nature; however, there were life-threatening conditions such as CA that highlight the need for detailed planning, multi-disciplined coordination, and communication to ensure a safe and secure event. In this environment, a detailed analysis of past data on appropriate responses to injured, sick, or CA runners is essential for operating safer marathon events in the future, and it is believed that sharing information and rescue manuals among all staff supporting the marathon, including volunteers, and strict adherence to them will contribute to the establishment of an appropriate medical rescue system, such as “All Yokohama.” Analysis of patient care, transport, and demographic data can assist in strengthening the knowledge and evidence to improve marathon medical and operational activities. Establishing this medical support system will contribute to safer marathon events and enable rapid measures and responses to health problems that occur during marathon races.

## Data Availability

All data produced in the present work are contained in the manuscript.

## Acknowledgements

We would like to thank all volunteer staff, Yokohama Fire Department, medical staff, and management staff of Peaceful Co., Ltd., who assisted with the entire medical support related to Yokohama Marathon. In addition, we would like to thank the hospital medical staff who accepted the injured and sick during the marathon. And we would like to thank Editage (www.editage.com) for English language editing.

## Declarations

We consulted with the Ethics Committee for research approval on this study, but it was determined that ethical approval was not required. So, ethical approval was not applicable.

## Consent for publication

Not applicable.

## Availability of data and materials

Data requests should be made to the corresponding authors.

## Informed Consent

Not applicable.

## Funding

This study did not receive any funding support.

## Competing interests

The authors declare that there are no conflicts of interest regarding the publication of this paper.

## Authors’ contributions

FO prepared the manuscript and collected the references. IT coordinated the authors. FO, RN, YN, YY, TK, KT, YF, SI and IT provided management support. KT helped to draft the manuscript. All authors have read and approved the final manuscript.

## Notes

### Competing Interest Statement

The authors have declared no competing interest.

### Author Declarations

Yokohama City University Hospital, Public University Corporation Medical and Hospital Management Department, Clinical Research Promotion Division, Ethics Office (1-1-1 Fukuura, Kanazawa-ku, Yokohama, Kanagawa 236-0004, Japan). After checking against internationally established research ethics guidelines, the judgement was determined that the study does not constitute "life science/medical research involving human subjects."

